# Spatial variation in housing construction material in low- and middle-income countries: a Bayesian spatial prediction model of a key infectious diseases risk factor and social determinant of health

**DOI:** 10.1101/2024.05.23.24307833

**Authors:** Josh M Colston, Bin Fang, Malena K Nong, Pavel Chernyavskiy, Navya Annapareddy, Venkataraman Lakshmi, Margaret N Kosek

## Abstract

Housing infrastructure and quality is a major determinant of infectious disease risk and other health outcomes in regions of the world where vector borne, waterborne and neglected tropical diseases are endemic. It is important to quantify the geographical distribution of improvements to the major dwelling components to identify and target resources towards populations at risk. The aim of this study was to model the sub-national spatial variation in housing materials using covariates with quasi-global coverage and use the resulting estimates to map the predicted coverage across the world’s low- and middle-income countries (LMICs). Data relating to the materials used in dwelling construction were sourced from nationally representative household surveys conducted since 2005. Materials used for construction of flooring, walls, and roof were reclassified as improved or unimproved. Households lacking location information were georeferenced using a novel methodology, and a suite of environmental and demographic spatial covariates were extracted at those locations for use as model predictors. Integrated nested Laplace approximation (INLA) models were fitted to obtain and map predicted probabilities for each dwelling component. The dataset compiled included information from households in 283,000 clusters from 350 surveys. Low coverage of improved housing was predicted across the Sahel and southern Sahara regions of Africa, much of inland Amazonia, and areas of the Tibetan plateau. Coverage of improved roofs and walls was high in the Central Asia, East Asia and Pacific and Latin America and the Caribbean regions, while improvements in all three components, but most notably floors, was low in Sub-Saharan Africa. Human development was by far the strongest determinant of dwelling component quality, though vegetation greenness and land use were also relevant markers These findings are made available to the reader as files that can be imported into a GIS for integration into relevant analysis to derive improved estimates of preventable health burdens attributed to housing.

## Introduction

The United Nations’ Sustainable Development Goals (SDGs) include ambitious commitments to fight communicable diseases (target 3.3) and provide adequate, safe and affordable housing (target 11.1) throughout its member states [1]. Although they fall under separate goals, housing quality has long been recognized as a social determinant of health and epidemiological evidence is now elucidating the mechanisms by which this relationship operates [2]. Many endemic infectious diseases of global public health concern, including several named in SDG3, are transmitted within and between households with the majority of infections occurring while the susceptible individual is at home [3], and consequently features of the built peridomestic environment and infrastructure play a role in promoting or impeding the spread of pathogens and their insect vectors [4]. This is particularly true of tropical and rural regions of Africa, Asia and Latin America where numerous vector borne and neglected tropical diseases circulate and where dwellings are often constructed using locally available, naturally occurring materials and traditional techniques such as wattle and daub, dried or burnt bricks, adobe, woven reed or bamboo and thatch [4]. These construction methods often require great skill and community mobilization to implement and are adapted over generations to suit local climate, ecology and topography, however numerous disease-causing insects and microbes are also well adapted to take advantage of the ecological niches that such buildings provide [5,6].

Infants and young children are particularly vulnerable to the health effects of housing construction material due to the high proportion of time spent in the family dwelling and behaviors common to early life such as crawling or playing on the floor [7–9]. Floors that are finished with wood, tiles or cement may protect against transmission of some diarrhea-causing enteric pathogens compared to those made of packed earth or sand either because they are easier to clean, or because they are less hospitable to pathogen survival outside the host [9].

As childhood mortality continues to decline globally, becoming concentrated in subnational hotspots it will be increasingly necessary to target interventions ever more specifically both geographically and to particular causes [10]. Several household-level determinants of health have been mapped at continental or global scale using survey data and spatial interpolation methods including water source and sanitation facility type [11], crowded living space [12], educational attainment [13], and relative wealth [14]. Tusting and colleagues have applied a similar approach to mapping houses built with finished materials across Sub-Saharan Africa for 2000 and 2015, defining such households as those having at least two out of three of the materials for the walls, roof and floor were finished, though they did not separate out these three components [15]. Building on these efforts, the aim of this study, a project of the Planetary Child Health & Enterics Observatory (Plan-EO, www.planeo.earth) [16] was to model the sub-national spatial variation in housing materials using covariates with quasi-global coverage and use the resulting estimates to map the predicted coverage across low- and middle-income countries (LMICs). The guiding hypothesis was that coverage of improved housing materials varies spatially as a function of environmental, and socio-demographic factors in a way that can be modelled using publicly available global datasets and state-of-the-art geostatistical methods.

## Materials and Methods

### Objective and scope

The objective of this analysis was to estimate the percent coverage of each category of materials used in dwelling component construction at all locations throughout the world’s LMICs (as defined by the Organisation for Economic Co-operation and Development [17], excluding those in Europe).

### Outcome variables

The categories of housing materials used in this analysis were those proposed by Florey and Taylor, who classify materials used for construction of flooring, walls, and roofs into natural, rudimentary, and finished types, and then further into improved and unimproved [18]. Data relating to these variables were compiled from nationally representative, population-based household surveys with two-stage cluster-randomized sample designs such as the Demographic and Health Surveys (DHS) [19], the Multiple Indicator Cluster Surveys (MICS) [20] and others. These programs collect information on coverage of health and development indicators and make the resulting microdata publicly available through their websites. All Standard DHSs, Malaria and AIDS Indicator Surveys (MIS and AIS) and MICS that collected information on housing material dating back to 2005 from all LMICs were included. For countries where no such surveys were available, either similar surveys from the 2000-2004 period or country-specific surveys were sourced where available. The unit of analysis was the household, and these were classified into three, mutually exclusive categories (natural, rudimentary, and finished) based on the housing material recorded by the survey interviewer for each of the three dwelling components (floors, walls, and roof) as shown in **Table 1**.

**Table 1:**
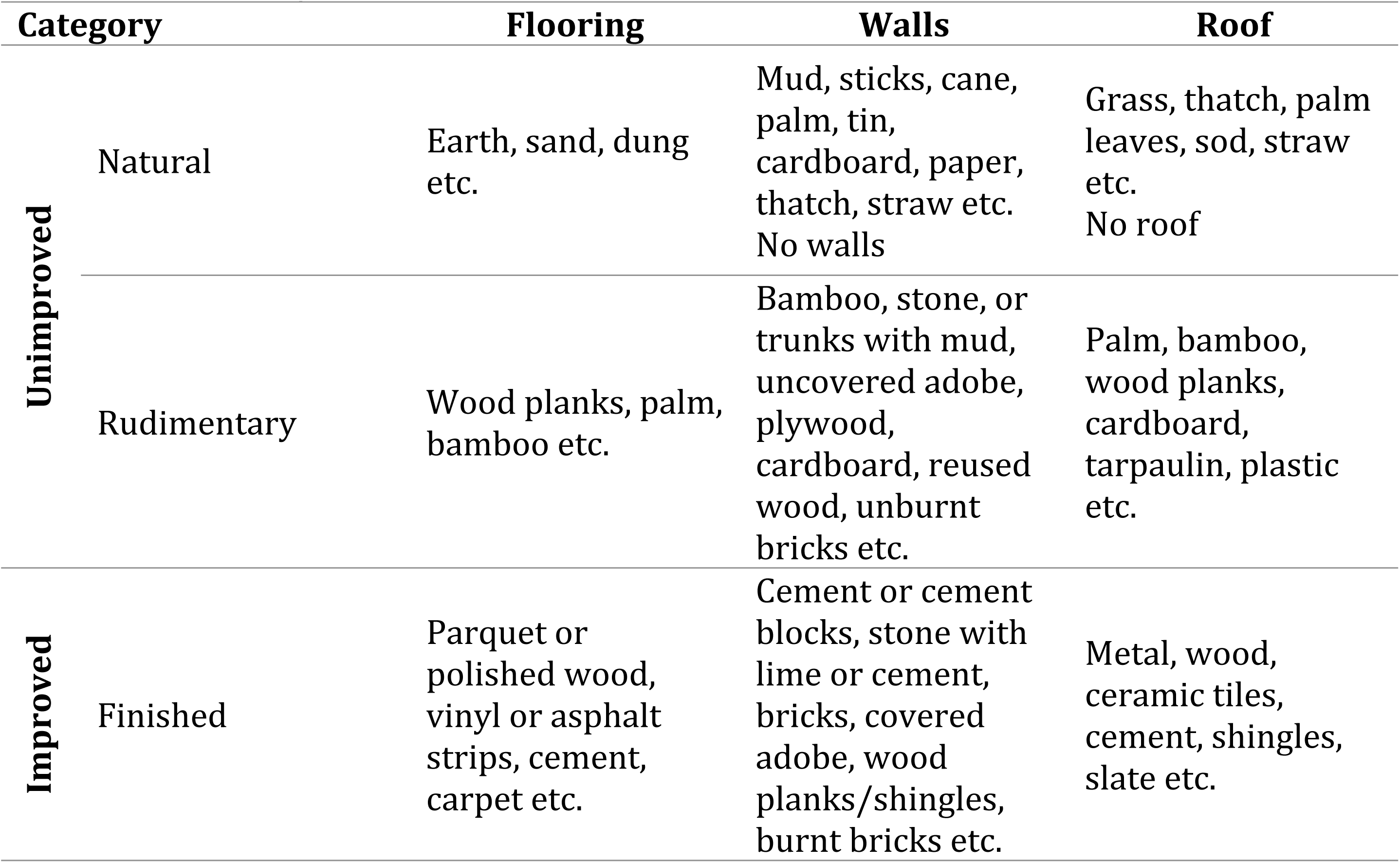
Classification of construction materials for the three components of the dwelling used as three-category outcome variables (adapted from Florey and Taylor 2016 [18])

### Georeferencing households

For this spatial analysis it was necessary to assign coordinates to each household representing its approximate location. Cluster-randomized surveys have a hierarchical design such that households are nested within clusters, the census enumeration areas that serve as the primary sampling unit, which are in turn nested within survey strata (sub-national region and urban/rural status). The DHS Program provides coordinates of the cluster centroids for most of the surveys they carry out [21] (though these are randomly “displaced” – systematically shifted up to a certain distance to preserve confidentiality [22]). However, these are not available for all clusters and surveys and equivalent coordinates have been made available only for a handful of MICS and no country-specific surveys. For this analysis, households were georeferenced to their displaced cluster centroid coordinates where available, otherwise their clusters were randomly assigned to populated settlement locations taken from the Humanitarian OpenStreetMap database [23] that fell within the same survey stratum (sub-national region and urban/rural status) with probability proportional to the population density of the settlement (extracted from the WorldPop [24] database at settlement coordinates). OpenStreetMap settlements were reclassified such that cities and towns were categorized as urban, and villages, hamlets, and isolated dwellings as rural. This novel cluster location assignment process was automated in ArcGIS Pro ModelBuilder [25] and Stata 18 [26].

### Covariates

A suite of time-static environmental and demographic spatial covariates available in raster format were compiled based on their hypothesized associations with the outcome variables. Definitions and sources of each covariate are shown in **Table 2**. Variable values were extracted at the georeferenced cluster locations in Python. In addition, time was calculated in continuous months since January 1^st^, 2005, based on the date of survey interview and log-transformed based on the assumption that changes in household material over time would be non-linear but unidirectional. Countries were grouped into the six regions used for administrative purposes by the World Bank [27], and this categorical variable was also treated as a covariate so that, for countries with no available survey data, estimates would be based partly on regional averages.

**Table 2:** Definitions and sources of variables included as covariate predictors in the model.

| Variable | Definition | Units/<br>Categories<sup>1</sup> | Source |
| --- | --- | --- | --- |
| **Accessibility to cities** | Travel time to nearest settlement of >50,000 inhabitants. | Minutes | MAP [33] |
| **Aridity index** | Mean annual precipitation / Mean annual reference evapotranspiration, 1970-2000. | Ratio | CGIAR-CSI [34] |
| **Climate zone** | First level Köppen-Geiger climate classification. | Tropical; arid; temperate; cold; polar | Beck et al. 2018 [35] |
| **Cropland areas** | Proportion of land given over to cropland, 2000. | Proportion | CIESIN [36] |
| **Distance to major river** | Distance to major perennial river (derived from rivers and lakes centerlines database). | Decimal degrees | Natural Earth [37] |
| **Elevation** | Elevation above sea level. | Meters | NOAA [38] |
| **Economic development** | Sub-national unit-level Gross Domestic Production (GDP) per capita, 2015 | Constant 2011 int. USD | Kummu et al. 2018 [39] |
| **Enhanced Vegetation Index** | Vegetation greenness corrected for atmospheric conditions and canopy background noise. | Ratio | USGS [40] |
| **Growing season length** | Reference length of annual agricultural growing period (baseline period 1961-1990). | Days | FAO, IIASA [41] |
| **Human development** | Sub-national unit-level Human Development Index (HDI), 2015 | Scale from 0 to 1 | Kummu et al. 2018 [39] |
| **Human Footprint Index** | Human Influence Index (HII) normalized by biome and realm. | Percentage | CIESIN [42] |
| **Irrigated areas** | Percentage of land equipped for irrigation around the year, 2000. | Percentage | FAO [43] |
| **Land cover and use** | General class of vegetation, tree, and ice cover or purpose of land use, 2020 (resampled and reclassified from Global Land Cover and Land Use) | Built up; cropland; desert; semi-arid; short vegetation; snow or ice; tree cover; wetland | GLAD [44] |
| **Land Surface Temperature** | Interannual averages of daily land surface temperature estimates for daytime, nighttime, and day/nighttime range, 2003-2020. | K | MOD21A1N v006 [45,46] |

| Variable | Definition | Units/<br>Categories<sup>1</sup> | Source |
| --- | --- | --- | --- |
| **Nighttime light** | The surface upward radiance from artificial light emissions extracted from at-sensor nighttime radiances at top-of-atmosphere. | nWatts·cm<sup>-2</sup>·sr<sup>-1</sup> | NASA Black Marble [47] |
| **Pasture areas** | Proportion of land given over to pasture, 2000. | Proportion | CIESIN [36] |
| **Population density** | Human population density per 1km<sup>2</sup>. | Inhabitants per km<sup>2</sup> | WorldPop [24] |
| **Potential evapotranspiration** | 8-day sum of the water vapor flux under ideal conditions of complete ground cover by plants. | kg/m<sup>2</sup>/8-day | NASA EOSDIS [48] |
| **Region** | Region of the globe as defined by the World Bank | East Asia & Pacific;<br>Europe & Central Asia;<br>Latin America & the Caribbean;<br>Middle East & North Africa;<br>South Asia | World Bank [27] |
| **Urbanicity** | Urbanicity status at georeferenced location (reclassified from Global Human Settlement database). | Urban; peri-urban; rural; remote | GHS [49] |

### Analysis

To reduce the database size and computational demands, and to neutralize the issue of within-cluster correlation, one household with non-missing outcome value was randomly sampled per cluster and retained for analysis (this selection was done separately for each of the three outcomes). Due to the computational demands of performing geospatial analysis at the global scale, we recoded all outcomes to be binary, by collapsing two of the response categories together (“rudimentary” was grouped with “natural”) to give “improved” / “unimproved” response categories as shown in **Table 1**, and in a modification of the schema proposed by Florey and Taylor (those authors grouped rudimentary and finished walls and roofs into the improved category, but not floors, however we opted for a consistent categorization across components to facilitate comparison between outcome variables [18]).

### Exploratory spatial data analysis

We first assessed the presence of spatial autocorrelation by generating semi-variograms of the Pearson residuals from a non-spatial logistic regression that included all explanatory variables listed in **Table 2** (Supplementary Figure S1). We fit spherical spatial correlation models to each semi-variogram and estimated the nugget, range, and sill for each outcome. The semi-variograms and respective models were estimated using the **gstat** R package [28]. Together with the nugget:sill ratio and the estimated range, we determined that an explicitly spatial modeling approach was required to account for the non-trivial spatial correlation in the Pearson residuals.

### Model fitting

Given the massive spatial scale of the database, with hundreds of thousands of points spanning most of the globe, incorporating spatial correlation into the models presented computational challenges. We used the **inlabru** R package to implement an integrated nested Laplace approximation (INLA) modeling approach in which all locations are projected onto a coarsened grid or “mesh” containing several thousand vertices that carry the spatial information and can be reprojected onto the observed data [29,30]. INLA models approximate Bayesian models by constructing the posterior distribution and then applying Laplace approximations, thus bypassing the need for time-consuming Markov chain Monte Carlo sampling and making global-scale computation feasible. All coordinates were transformed via the Mollweide projection and scaled into kilometers prior to analysis. The mesh used for modelling had 18,352 vertices, placed within continental boundaries. Further details on the implementation of the INLA model are provided in **Supplementary File 1**.

### Model predictions

Predicted probabilities for each outcome were made for all locations in the domain of interest (the LMICs) at 5 km^2^ resolution and exported in Georeferenced Tag Image File format (GeoTIFF). The spatial covariates from Table 2 along with the time variable were used to generate predicted logistic distribution probability of the finished class of each building material from the INLA model. A value for time corresponding to the first of January 2023 was used for making predictions. Missing pixel values were filled by performing imputation using k-Nearest Neighbors method by Python Scikit-learn package [31].

### Model evaluation

The predictive performance of the spatial models was assessed by calculating common metrics of recall (sensitivity), precision (positive predictive value), accuracy (the proportion correctly classified), F1-score (mean of precision and recall), and area under the receiver operating characteristic curve (ROC-AUC). For each performance metric, two multiclass averaging metrics (macro and weighted average) were calculated, including macro averaging and weighted macro averaging, given by:

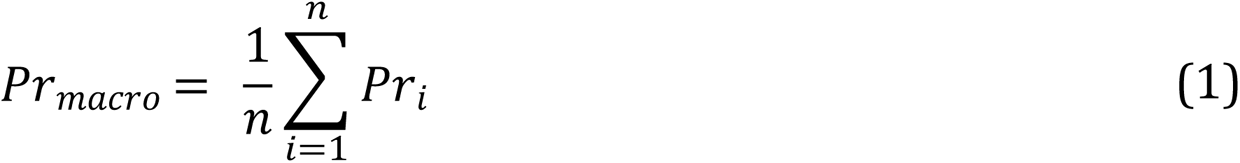

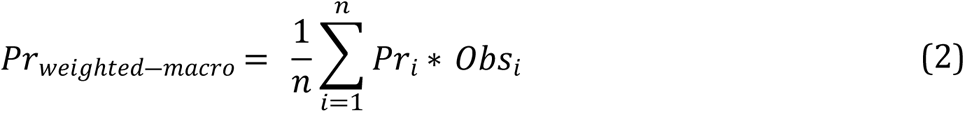

Where *Pr*_*i*_ is the precision calculated from the multiple class predictions and *Obs*_*i*_ is the number of observations of one class. *n* is the total number of observations of all classes. To assess the relative contribution of each covariate to the models, feature importance values for the input raster covariates were calculated by running parallel non-spatial linear regression models (since the **inlabru** package does not provide feature importance output) that were otherwise identically specified and scaling the output coefficients to the 0 – 1 range using the Scikit-learn Python package. These feature importance values can be interpreted as conditional associations, quantifying the responded variation of the output when only the given feature is allowed to vary while all other features are held constant.

### Ethics statement

All human subject information used in this analysis was anonymized, publicly available secondary data, and therefore ethical approval was not required or sought. For data provided by the DHS Program, data access requests (including for the displaced cluster coordinates) were submitted and authorized through the Program’s website. A completed checklist of Guidelines for Accurate and Transparent Health Estimates Reporting (GATHER [32]) is included in **Supplementary File S1**.

## Results

350 nationally representative household surveys (together containing data from more than 6 million households in 283,000 clusters) met the inclusion criteria, reported information on construction material types for one or more of the dwelling components and were included in the model training dataset. **Figure 1** shows the number of surveys contributed by each LMIC, while **Supplementary File S2** gives the national level distribution of each of the three housing construction variables in each survey (before within-cluster sub-sampling, and without sample weights applied). All eligible surveys included information on floor material; however, wall and roof material information were only available from 328 and 324 surveys respectively. No relevant data from household surveys could be found for several LMICs with large geographies and populations, most notably China, Iran, Venezuela, Libya, and Malaysia, as well as the smaller countries of Eritrea, North Korea, Lebanon, Equatorial Guinea, and numerous island nations such as Sri Lanka.

**Figure 1:**
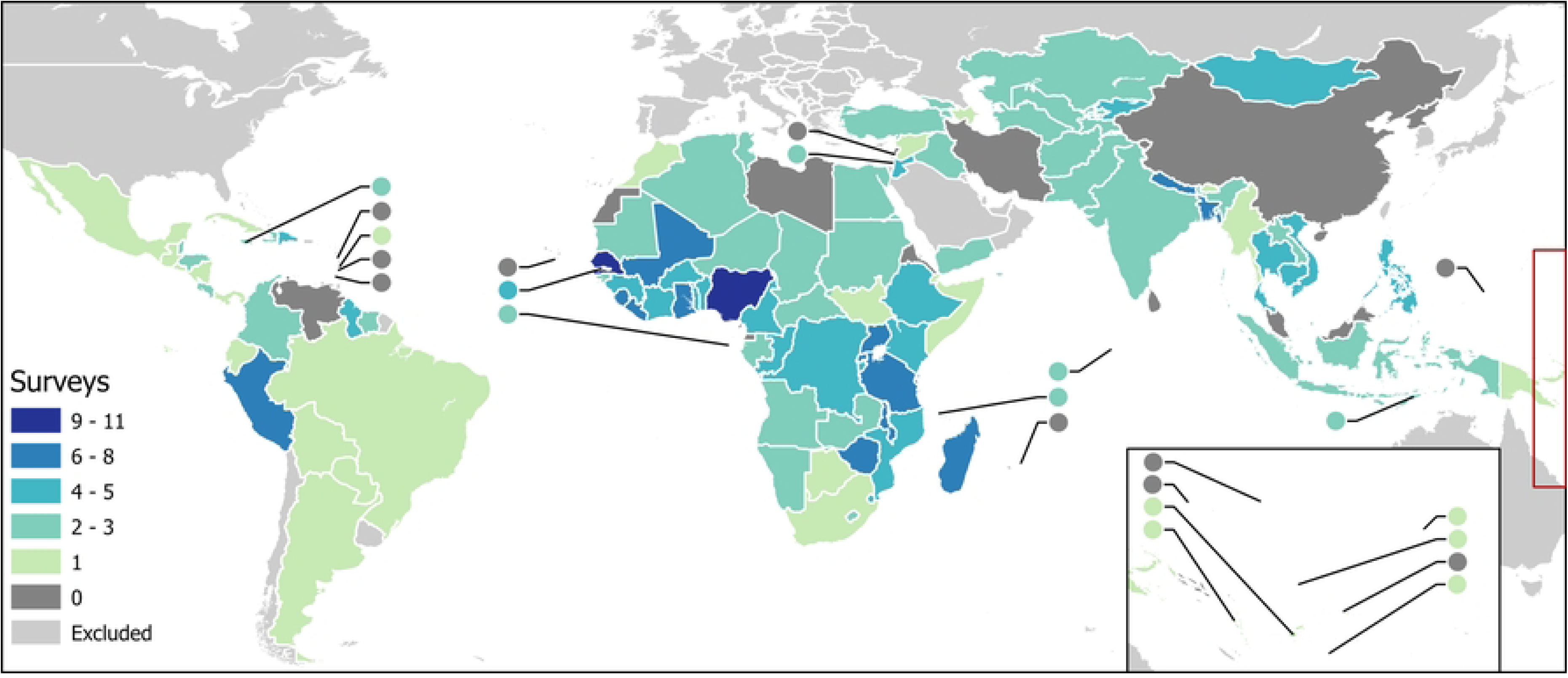
Number of nationally representative household surveys included in input dataset by country for included LMICs (small countries represented by circles). Base map compiled from shapefiles obtained from U.S. Department of State—Humanitarian Information Unit [50] and Natural Earth free vector map data @ naturalearthdata.com that are made available in the public domain with no restrictions.

Figure 2 shows the geographical distribution of the coverage of improved materials predicted by the INLA models for each of the three binary dwelling component variables across the domain of included LMICs. These predictions are also provided as raster TIFF files available on the Dryad data repository. There are some similarities across the variables, with low coverage predicted for all three across a wide belt of the Sahel and southern Sahara regions of Africa, much of inland Amazonia, and areas of the Tibetan plateau, as well as individual countries including the Democratic Republic of the Congo, Mozambique, Madagascar, Pakistan, and Papua New Guinea. High coverage of all three improved components coincided across much of the Middle East, Mediterranean North Africa, the coast of the Bight of Benin, the Caribbean, sub-Amazonian Brazil, southern Argentina, and South Africa. However, divergence in coverage of the three variables is evident across many locations. Across Kazakhstan, Mongolia, Azerbaijan, Cambodia and Laos, low coverage of improved floors, but high coverage of walls and roofs were predicted, while in Afghanistan, the reverse was the case. Yemen has mostly high improved floor coverage predicted, but low improved roof and mixed improved wall coverage, while on the island of Borneo, that pattern is reversed. Importantly, sub-national patterns are clearly visible, for example, with respect to improved floors, walls, and roofs in India, China, Mexico, and Brazil.

**Figure 2:**
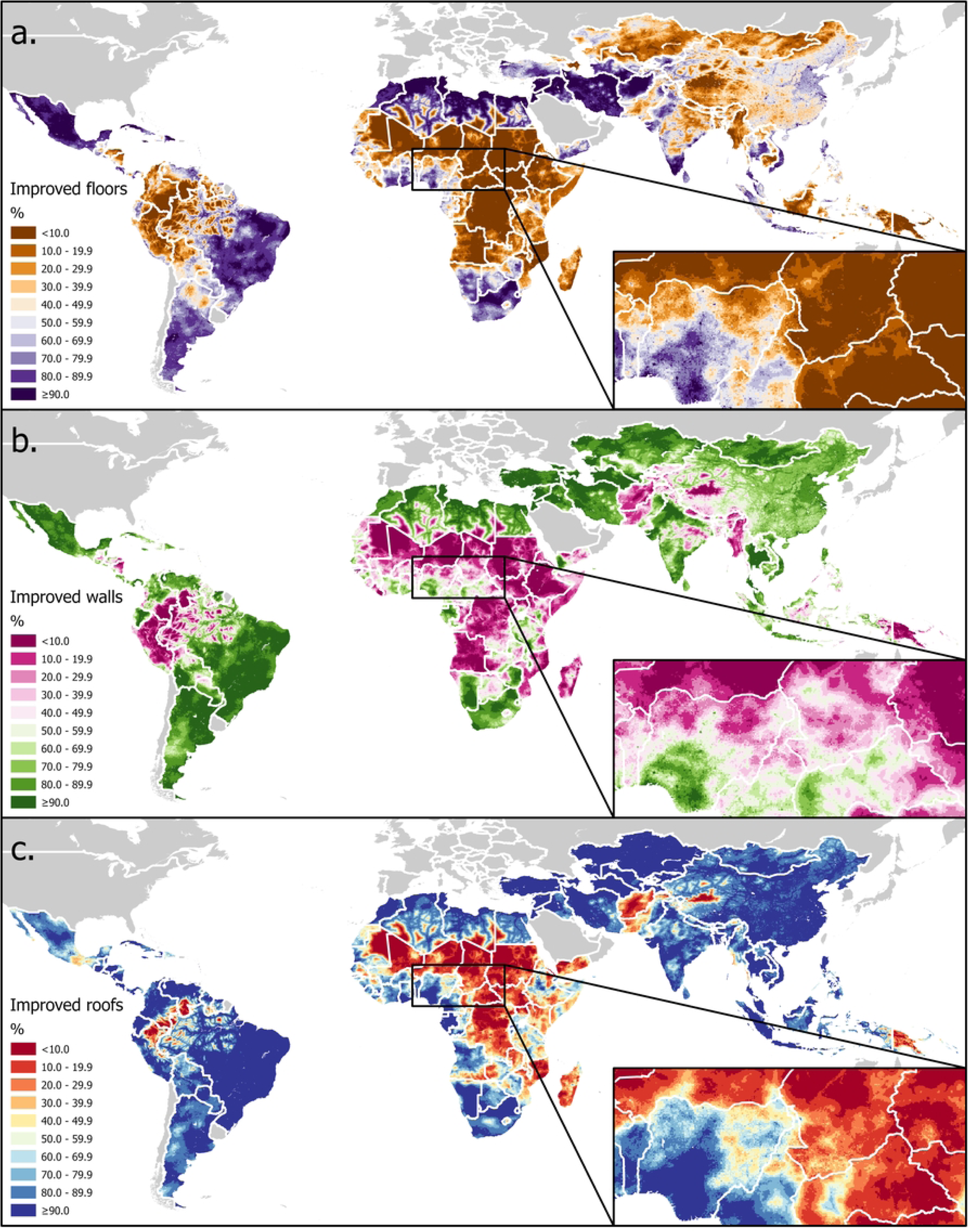
Coverage of improved material for three dwelling components - a. floors, b. walls, c. roofs – in LMICs predicted by integrated nested Laplace approximation (INLA) models fitted to household survey data. Base maps compiled from shapefiles obtained from U.S. Department of State—Humanitarian Information Unit [50] and Natural Earth free vector map data @ naturalearthdata.com that are made available in the public domain with no restrictions.

Figure 3 shows ridge plots visualizing the distribution of predicted values for the coverage of improved status for each of the three dwelling components and stratified by the six world regions. The distribution of improved roofs was highly concentrated at values very close to 100% in the Central Asia region, findings which are borne out by the input data, in which most surveys recorded a coverage of finished roofs greater than 97% (**Supplementary File S2**). This was true to a far lesser extent for other regions - with the exception of Sub-Saharan Africa, which had predicted values much more evenly dispersed along the range of values – and for improved walls, though the South Asia region and had a much more dispersed, bimodal distribution for the latter variable. For improved floors, predicted values were highly concentrated at the low extreme of Sub-Saharan Africa.

**Figure 3:**
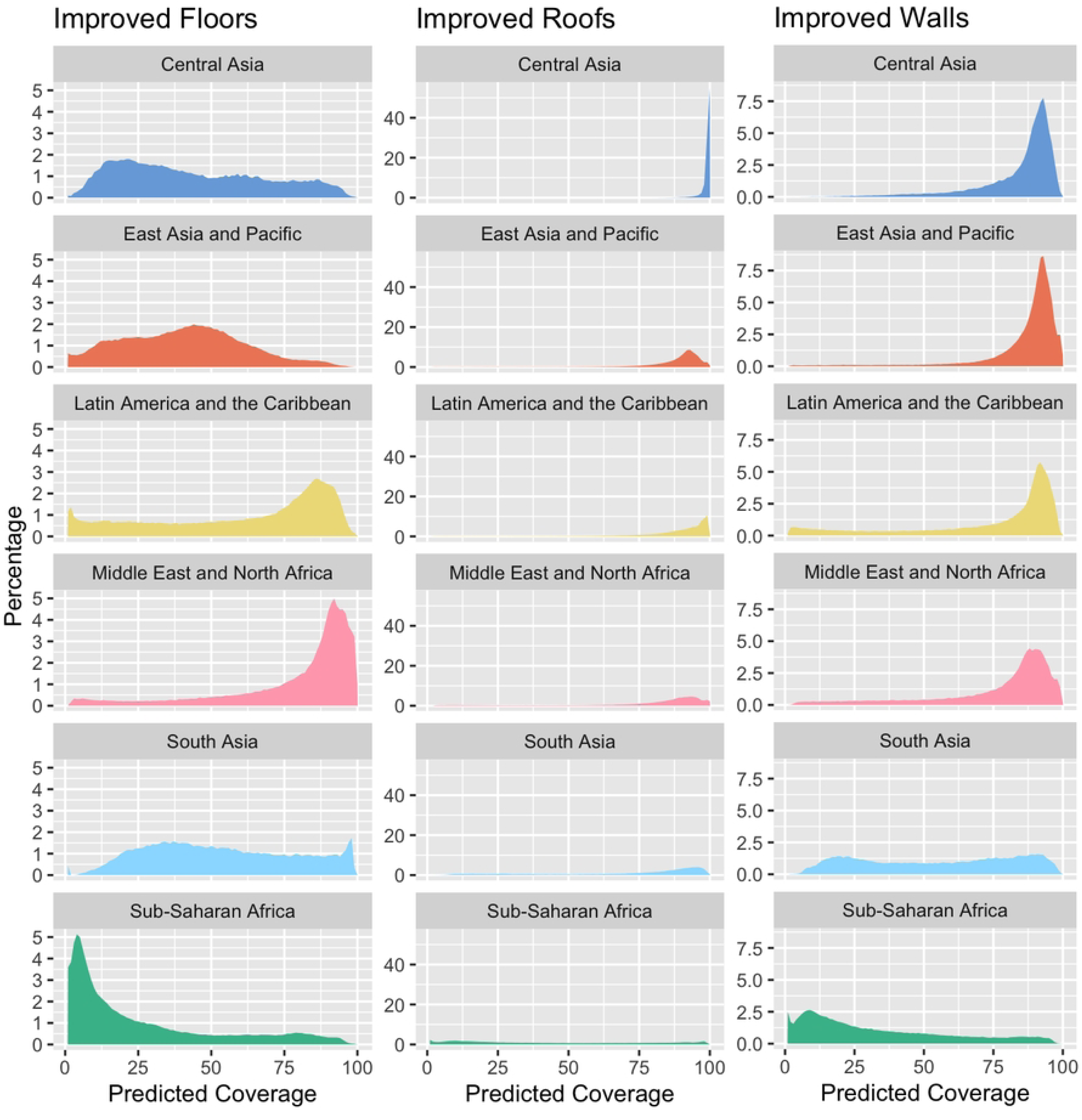
Distribution of values predicted for coverage of improved dwelling components by INLA models, stratified by component and world region.

Figure 4 visualizes the feature importance values for each covariate in each of the three models. More than half (eleven) of the variables did not contribute to any of the models. Feature importance was dominated by the same single variable (human development index), accounting for more than 50% of the variation in all three models. For the walls and to a lesser extent the floors models, the next most important feature was provided by the enhanced vegetation index (EVI), whereas for the roofs model, cropland and pasture areas contributed more to the model prediction, with EVI ranking fourth.

**Figure 4:**
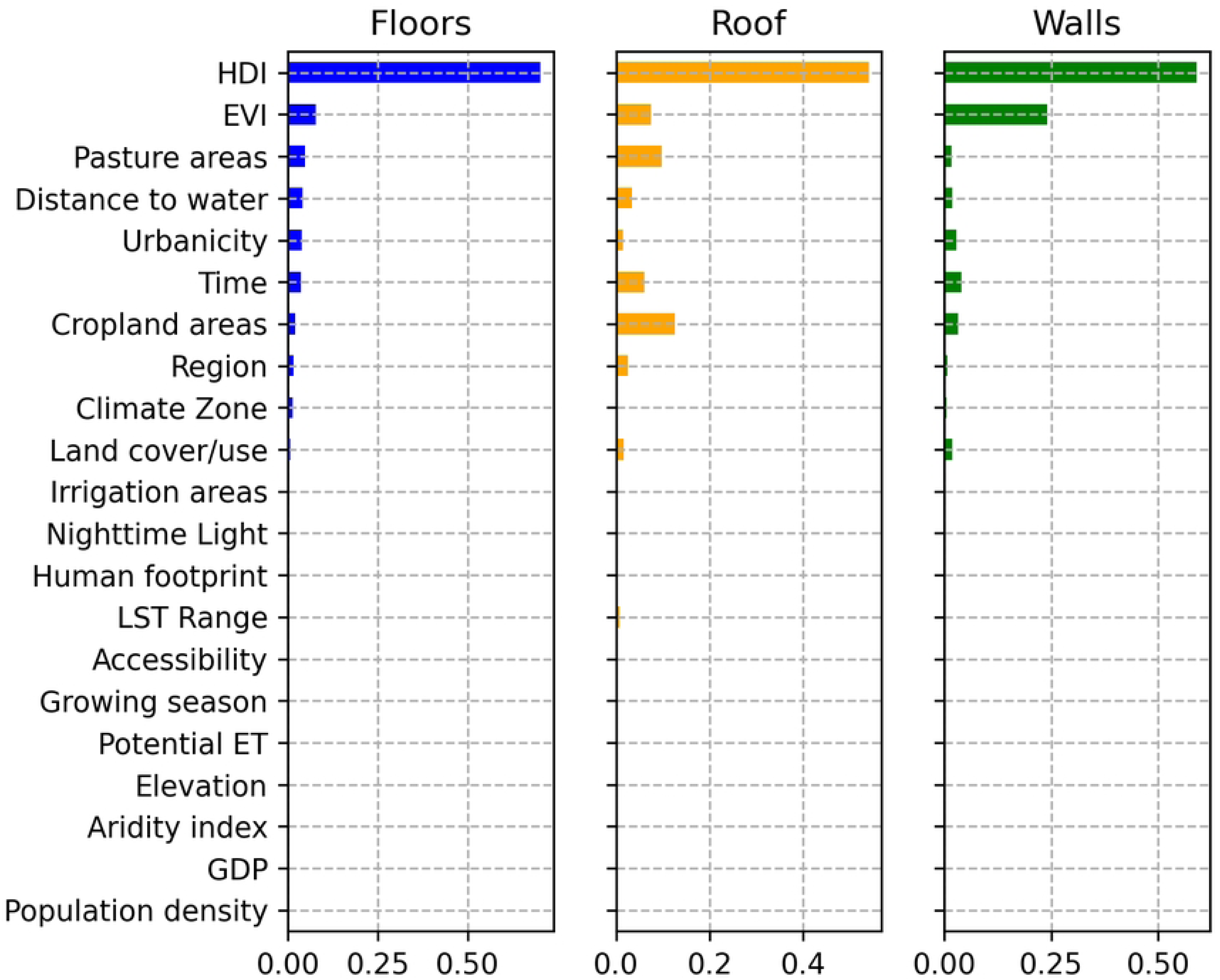
Feature importance for each covariate included in the final model for each of the dwelling components (HDI – Human Development Index; EVI – Enhanced Vegetation Index; LST – Land Surface Temperature; ET – Evapotranspiration; GDP – Gross Domestic Product).

**Table 3** gives statistics that evaluate the models’ performance in classifying household construction material types for the three dwelling components. Across the whole database, floors were the dwelling component for which coverage of improved construction material was lowest at 57.9%, the equivalent coverage for walls and roofs being 67.1% and 80.3% respectively. While precision, recall and F1-score statistics were generally high for the unimproved category in all models, they varied considerably for the improved category, particularly for the roofs model, for which recall, and F1-score were just 0.4 and 0.5 respectively. However, the roofs model was the one with the highest weighted average for those three statistics (a precision of 0.84, recall of 0.85 and F1-score of 0.83, compared with 0.78, 0.79, and 0.78 respectively for the walls and 0.77 for all three statistics for the floors model). All three models demonstrated similarly strong discriminatory power and performance in distinguishing between households with improved and unimproved construction materials in the respective dwelling components, with ROC-AUC statistics of 0.85 – 0.87.

**Table 3:** Evaluation statistics for models of construction materials for three dwelling components.

|  |  | **Observations (%)** | **Precision** | **Recall** | **F1-score** | **ROC-AUC** |
| --- | --- | --- | --- | --- | --- | --- |
| **Floors** | **Total** | 258,472 (100.0) | - | - | - | 0.85 |
|  | **Unimproved** | 108,931 (42.1) | 0.73 | 0.74 | 0.73 | - |
|  | **Improved** | 149,541 (57.9) | 0.81 | 0.80 | 0.80 | - |
|  | **Macro-average** | - | 0.77 | 0.77 | 0.77 | - |
|  | **Weighted average** | - | 0.77 | 0.77 | 0.77 | - |
| **Walls** | **Total** | 248,421 (100.0) | - | - | - | 0.85 |
|  | **Unimproved** | 81,621 (32.9) | 0.71 | 0.59 | 0.65 | - |
|  | **Improved** | 166,800 (67.1) | 0.82 | 0.88 | 0.85 | - |
|  | **Macro-average** | - | 0.77 | 0.74 | 0.75 | - |
|  | **Weighted average** | - | 0.78 | 0.79 | 0.78 | - |
| **Roofs** | **Total** | 235,024 (100.0) | - | - | - | 0.87 |
|  | **Unimproved** | 46,272 (19.7) | 0.76 | 0.38 | 0.50 | - |
|  | **Improved** | 188,752 (80.3) | 0.86 | 0.97 | 0.91 | - |
|  | **Macro-average** | - | 0.81 | 0.67 | 0.71 | - |
|  | **Weighted average** | - | 0.84 | 0.85 | 0.83 | - |

## Discussion

Housing infrastructure and quality are major determinants of infectious disease risk and other health outcomes, particularly in regions of the world where vector borne, waterborne and neglected tropical diseases are endemic. Although, the nature of this relationship is complex and multifaceted and varies depending on the specific pathogen and vector species, it highlights the importance of targeting interventions to mitigate these adverse health outcomes, particularly in LMICs where the overwhelming majority of childhood mortality occurs. As attention turns to improving housing quality in low-resource settings as a strategy for controlling infectious diseases, it is important to quantify the geographical distribution of improvements to the major dwelling components to identify and target resources towards populations at risk. This study is the first attempt to meet this objective.

The importance of housing materials is clearly not restricted to vectorborne diseases. Finished floors have been associated with decreases of 0.89 in Log_10_ *E. coli* contamination in Peru [51], 78% in intestinal parasite prevalence in Mexican children [52], and 9% for diarrheal disease risk, 11% for both enteric bacteria and enteric protozoa risk [8], and 17% for *Shigella* spp. infection probability in meta-analyses of children under 5 years across multiple LMIC surveillance sites [53]. Traditional roof material has also been shown to be associated with childhood diarrhea [54], even after adjusting for floor material [55]. Pooled analyses of household survey data from multiple countries have found associations of living in improved housing on numerous child health outcomes, including cognitive and social-emotional development [7], and nutritional status [56], in addition to malaria infection [18,57]. Additionally, there is evidence of increased acute respiratory illness (ARI) in children in Pakistan, with unimproved flooring increasing ARI risk by 18%, and unimproved walling materials also increasing the risk of ARI in children under the age of five [58]. These findings are supported by similar findings with different studies in India, Nigeria, ad Lao PDR [59–61].

This study is subject to several limitations. Our characterization of housing was constrained by the availability of data from household surveys, which generally only ask about just three components, and don’t include questions about other relevant features of the built household environment, such as screens covering openings [62] elevation of sleeping areas or improvements to windows and ventilation [63]. Although the variables were originally in three-class ordinal categorical format, we had to combine categories and model them as dichotomous, because there is currently no way to address adjacent categories and parallel odds using the INLA modeling approach. Additionally, our spatial models assume a stationary (i.e., global) covariance structure that does not vary across the globe. This is likely an oversimplification of the latent spatial effects; however, estimating a non-stationary spatial model at the global scale falls outside the scope of the current article and presents a worthwhile future direction. Likewise, improving the precision of the mesh used by INLA may improve predictions, but with ROC-AUC values already relatively high, this is likely to yield only marginal gains.

Despite these limitations, the product developed fills an important gap in spatially characterizing determinants of the principal causes of infectious disease burden in LMICs. Many types of mosquitoes such as those that transmit malaria (*Anopheles* spp.), dengue (*Aedes* spp.), filariasis and Japanese encephalitis (*Culex* spp.) enter the home through eaves and other openings [64] and rest on walls and ceilings after ingesting a blood meal (the basis behind indoor residual spraying [IRS] of these surfaces as a malaria control intervention). Indeed, in Africa, 80% of malaria transmission occurs indoors [3] and houses with roofs and walls constructed of natural material provide more points of entry [64,65] and preferred resting places [66] for malaria-transmitting mosquitoes, insights which are increasingly putting housing improvements on the research agendas as potential disease control strategies [63,65]. In rural Gambia, studies have found reductions in intradomiciliary mosquito vector abundance and survival through installing plywood ceilings [67], closing eaves in thatched roofs [68,69], and replacing thatch with ventilated metal roofing [70]. In rural Uganda, living in a house constructed of traditional materials (thatched roof, mud walls, earth floor etc.) has been associated with increased clinical malaria incidence [71] and parasitemia in children [72] and pregnant women [73], and decreased effectiveness of IRS in reducing *Anopheles* biting rates [72]. Similar protective effects of improved housing construction material on entomological and clinical malaria outcomes have been documented separately in Burkina Faso [74], Ethiopia [75], Laos [76], Malawi [77], South Africa [78], and Tanzania [79], while pooled effects from systematic reviews have been reported on the order of a 32% reduction in mosquito-borne diseases, 47% for malaria infection and 85% for indoor vector densities [65,80]. Aside from mosquito-borne illnesses, living in households with walls made of mud or thatch carries an increased risk of leishmaniasis infection and indoor abundance of sandfly vectors [81], while in the Americas, Chagas Disease vectors (triatomine bugs) are drawn to houses with thatched palm roofs and mud walls [82]. In a Guatemalan community, for example, the odds of triatomine presence were 3.85 times higher in houses with walls that lacked plastering [83], while in rural Paraguay, an intervention to provide houses with smooth, flat and crack-free walls, reduced triatomine infestation by 96.4%, a comparable effect to that of fumigation [84].

## Conclusions

In conclusion, this study applies a relatively computationally efficient and spatially explicit modeling approach to a very large dataset, representative of but standardized across diverse geographies, and collected through rigorous and standardized methodologies. The findings allow us to assess the predictive performance of the models as well as the relative contribution of particular covariate variables, and the resulting predictions are made available to the reader in a readily useable format (available from www.datadryad/org). Human development is by far the strongest determinant of dwelling component quality, though vegetation greenness and land use (cropland and pasture) are also relevant markers. Prevalence of improved roofs and walls is high in the Central Asia, East Asia and Pacific and Latin America and the Caribbean regions, while coverage of improvements in all three components, but most notably floors, is low in Sub-Saharan Africa.

## Data Availability

The data used in this analysis are publicly available from the sources listed in table 2 and supplementary file S2. The statistical source code used to generate estimates can be accessed is available from the corresponding author upon reasonable request.

## References

1. United Nations. 17 Goals - Learn About the SDGs. [cited 26 Oct 2018]. Available: http://17goals.org/

2. Krieger J, Higgins DL. Housing and health: time again for public health action. Am J Public Health. 2002;92: 758–768. doi:10.2105/ajph.92.5.758

3. Haines A, Bruce N, Cairncross S, Davies M, Greenland K, Hiscox A, et al. Promoting health and advancing development through improved housing in low-income settings. J Urban Health. 2013;90: 810–831. doi:10.1007/s11524-012-9773-8

4. Knudsen J, von Seidlein L. Healthy homes in tropical zones: improving rural housing in Asia and Africa. London: Axel Menges; 2014.

5. Costantini C, Sagnon N, della Torre A, Coluzzi M. Mosquito behavioural aspects of vector-human interactions in the Anopheles gambiae complex. Parassitologia. 1999;41: 209–217.

6. Trpis M, Hausermann W. Genetics of house-entering behaviour in East African populations of Aedes aegypti (L.) (Diptera: Culicidae) and its relevance to speciation. Bulletin of Entomological Research. 1978;68: 521–532. doi:10.1017/S0007485300009494

7. Gao Y, Zhang L, Kc A, Wang Y, Zou S, Chen C, et al. Housing environment and early childhood development in sub-Saharan Africa: A cross-sectional analysis. PLoS Med. 2021;18: e1003578. doi:10.1371/journal.pmed.1003578

8. Colston JM, Faruque ASG, Hossain MJ, Saha D, Kanungo S, Mandomando I, et al. Associations between Household-Level Exposures and All-Cause Diarrhea and Pathogen-Specific Enteric Infections in Children Enrolled in Five Sentinel Surveillance Studies. International Journal of Environmental Research and Public Health. 2020;17: 8078. doi:10.3390/ijerph17218078

9. Schiaffino F, Rengifo Trigoso D, Colston JM, Paredes Olortegui M, Shapiama Lopez WV, Garcia Bardales PF, et al. Associations among Household Animal Ownership, Infrastructure, and Hygiene Characteristics with Source Attribution of Household Fecal Contamination in Peri-Urban Communities of Iquitos, Peru. Am J Trop Med Hyg. 2021;104: 372–381. doi:10.4269/ajtmh.20-0810

10. Perin J, Mulick A, Yeung D, Villavicencio F, Lopez G, Strong KL, et al. Global, regional, and national causes of under-5 mortality in 2000–19: an updated systematic analysis with implications for the Sustainable Development Goals. The Lancet Child & Adolescent Health. 2022;6: 106–115. doi:10.1016/S2352-4642(21)00311-4

11. Deshpande A, Miller-Petrie MK, Lindstedt PA, Baumann MM, Johnson KB, Blacker BF, et al. Mapping geographical inequalities in access to drinking water and sanitation facilities in low-income and middle-income countries, 2000–17. The Lancet Global Health. 2020;8: e1162–e1185. doi:10.1016/S2214-109X(20)30278-3

12. Chipeta MG, Kumaran EPA, Browne AJ, Hamadani BHK, Haines-Woodhouse G, Sartorius B, et al. Mapping local variation in household overcrowding across Africa from 2000 to 2018: a modelling study. The Lancet Planetary Health. 2022;6: e670– e681. doi:10.1016/S2542-5196(22)00149-8

13. Graetz N, Woyczynski L, Wilson KF, Hall JB, Abate KH, Abd-Allah F, et al. Mapping disparities in education across low- and middle-income countries. Nature. 2019. doi:10.1038/s41586-019-1872-1

14. Data For Good at Meta. Relative Wealth Index. 2022 [cited 2 Sep 2022]. Available: https://dataforgood.facebook.com/dfg/tools/relative-wealth-index#methodology

15. Tusting LS, Bisanzio D, Alabaster G, Cameron E, Cibulskis R, Davies M, et al. Mapping changes in housing in sub-Saharan Africa from 2000 to 2015. Nature. 2019 [cited 17 Oct 2019]. doi:10.1038/s41586-019-1050-5

16. Colston JM, Chernyavskiy P, Gardner LM, Fang B, Houpt E, Swarup S, et al. The Planetary Child Health and Enterics Observatory (Plan-EO): a Protocol for an Interdisciplinary Research Initiative and Web-Based Dashboard for Climate-Informed Mapping of Enteric Infectious Diseases and their Risk Factors and Interventions in Low- and Middle-Income Countries. 18 Apr 2023 [cited 24 Apr 2023]. doi:10.21203/rs.3.rs-2640564/v2

17. Organisation for Economic Co-operation and Development. DAC List of ODA Recipients. In: OECD [Internet]. 2020 [cited 10 Dec 2021]. Available: https://www.oecd.org/dac/financing-sustainable-development/development-finance-standards/daclist.htm

18. Florey L, Taylor C. Using household survey data to explore the effects of improved housing conditions on malaria infection in children in Sub-Saharan Africa. Rockville, Maryland, USA: ICF International; 2016 Aug. Available: https://dhsprogram.com/publications/publication-AS61-Analytical-Studies.cfm

19. ICF International. Demographic and Health Surveys (various, 2000-2021). Rockville, Maryland, USA: ICF International; 2021.

20. UNICEF. Multiple Indicator Cluster Surveys (various, 2000-2021). New York, NY: UNICEF; 2021.

21. Perez-Haydrich C, Warren JL, Burgert CR, Emch ME. Guidelines on the use of DHS GPS data. 2013 [cited 22 Jul 2022]. Available: https://dhsprogram.com/publications/publication-SAR8-Spatial-Analysis-Reports.cfm

22. Burgert C, Colston J, Roy T, Zachary B. Geographic displacement procedure and georeferenced data release policy for the Demographic and Health Surveys. Calverton, MD, USA: MEASURE DHS; 2013 Sep. Available: http://dhsprogram.com/publications/publication-SAR7-Spatial-Analysis-Reports.cfm

23. OpenStreetMap© contributors. Humanitarian OpenStreetMap Populated Places Datasets (OpenStreetMap Export). 2022 [cited 22 Jul 2022]. Available: https://data.humdata.org/organization/hot

24. Tatem AJ. WorldPop, open data for spatial demography. Scientific Data. 2017;4: 170004. doi:10.1038/sdata.2017.4

25. ESRI. ArcGIS Desktop: Release 10.8. Redlands, CA: Environmental Systems Research Institute; 2019. Available: https://desktop.arcgis.com/en/desktop/

26. StataCorp. Stata Statistical Software: Release 18. College Station, TX: StataCorp LLC; 2023.

27. The World Bank. The world by region. In: SDG Atlas 2017 [Internet]. 2017 [cited 10 Dec 2021]. Available: https://datatopics.worldbank.org/sdgatlas/archive/2017/the-world-by-region.html

28. Pebesma EJ. Multivariable geostatistics in S: the gstat package. Computers & Geosciences. 2004;30: 683–691. doi:10.1016/j.cageo.2004.03.012

29. Bachl FE, Lindgren F, Borchers DL, Illian JB. inlabru: an R package for Bayesian spatial modelling from ecological survey data. Methods in Ecology and Evolution. 2019;10: 760–766. doi:10.1111/2041-210X.13168

30. Lindgren F, Bachl FE, Borchers DL, Simpson D, Scott-Howard L, Andy S, et al. inlabru: Bayesian Latent Gaussian Modelling using INLA and Extensions. 2023. Available: https://cran.r-project.org/web/packages/inlabru/index.html

31. Pedregosa F, Varoquaux G, Gramfort A, Michel V, Thirion B, Grisel O, et al. Scikit-learn: Machine Learning in Python. Journal of Machine Learning Research. 2011;12: 2825– 2830.

32. Stevens GA, Alkema L, Black RE, Boerma JT, Collins GS, Ezzati M, et al. Guidelines for Accurate and Transparent Health Estimates Reporting: the GATHER statement. The Lancet. 2016;388: e19–e23. doi:10.1016/S0140-6736(16)30388-9

33. Weiss DJ, Nelson A, Gibson HS, Temperley W, Peedell S, Lieber A, et al. A global map of travel time to cities to assess inequalities in accessibility in 2015. Nature. 2018;553: 333–336. doi:10.1038/nature25181

34. Trabucco A, Zomer R J. Global Aridity Index and Potential Evapo-Transpiration (ET0). CGIAR Consortium for Spatial Information (CGIAR-SCI); 2018. Report No.: 2. Available: https://cgiarcsi.comunity

35. Beck HE, Zimmermann NE, McVicar TR, Vergopolan N, Berg A, Wood EF. Present and future Köppen-Geiger climate classification maps at 1-km resolution. Sci Data. 2018;5: 180214. doi:10.1038/sdata.2018.214

36. Ramankutty N, Evan AT, Monfreda C, Foley JA. Farming the planet: 1. Geographic distribution of global agricultural lands in the year 2000. Global Biogeochemical Cycles. 2008;22: n/a–n/a. doi:10.1029/2007GB002952

37. Natural Earth. Rivers and Lakes Centerlines 4.1.0. 2021. Available: https://www.naturalearthdata.com/downloads/10m-physical-vectors/10m-rivers-lake-centerlines/

38. Hastings DA, Dunbar PK. Global Land One-kilometer Base Elevation (GLOBE) Digital Elevation Model, Documentation, Volume 1.0. Boulder, Colorado: National Oceanic and Atmospheric Administration, National Geophysical Data Center; 1999.

39. Kummu M, Taka M, Guillaume JHA. Gridded global datasets for Gross Domestic Product and Human Development Index over 1990–2015. Sci Data. 2018;5: 180004. doi:10.1038/sdata.2018.4

40. U.S. Geological Survey. Landsat Enhanced Vegetation Index. In: Landsat Missions [Internet]. 2021 [cited 14 Dec 2021]. Available: https://www.usgs.gov/landsat-missions/landsat-enhanced-vegetation-index

41. The Food and Agriculture Organization (FAO), International Institute of Applied Systems Analysis. Global Agro-ecological Zones (GAEZ v3.0). Rome, Italy and Laxenburg, Austria: FAO & IIASA; 2012.

42. Wildlife Conservation Society, Center for International Earth Science Information Network - CIESIN. Last of the Wild Project, Version 2, 2005 (LWP-2): Global Human Footprint Dataset (Geographic). Palisades, NY: NASA Socioeconomic Data and Applications Center (SEDAC); 2005. Available: https://sedac.ciesin.columbia.edu/data/set/wildareas-v2-human-footprint-geographic

43. Siebert S, Döll P, Hoogeveen J, Faures J-M, Frenken K, Feick S. Development and validation of the global map of irrigation areas. Hydrology and Earth System Sciences. 2005;9: 535–547. doi:10.5194/hess-9-535-2005

44. Potapov P, Hansen MC, Pickens A, Hernandez-Serna A, Tyukavina A, Turubanova S, et al. The Global 2000-2020 Land Cover and Land Use Change Dataset Derived From the Landsat Archive: First Results. Front Remote Sens. 2022;3. doi:10.3389/frsen.2022.856903

45. Hulley, Glynn, Hook, Simon. MOD21A1D MODIS/Terra Land Surface Temperature/3-Band Emissivity Daily L3 Global 1km SIN Grid Day V006. NASA EOSDIS Land Processes DAAC; 2017. doi:10.5067/MODIS/MOD21A1D.006

46. Hulley, Glynn, Hook, Simon. MOD21A1N MODIS/Terra Land Surface Temperature/3-Band Emissivity Daily L3 Global 1km SIN Grid Night V006. NASA EOSDIS Land Processes DAAC; 2017. doi:10.5067/MODIS/MOD21A1N.006

47. Román MO, Wang Z, Sun Q, Kalb V, Miller SD, Molthan A, et al. NASA’s Black Marble nighttime lights product suite. Remote Sensing of Environment. 2018;210: 113–143. doi:10.1016/j.rse.2018.03.017

48. Running, Steve, Mu, Qiaozhen, Zhao, Maosheng. MOD16A2 MODIS/Terra Net Evapotranspiration 8-Day L4 Global 500m SIN Grid V006. NASA EOSDIS Land Processes DAAC; 2017. doi:10.5067/MODIS/MOD16A2.006

49. Pesaresi M, Ehrlich D, Stefano F, Florcyk A, Freire SMC, Halkia S, et al. Operating procedure for the production of the Global Human Settlement Layer from Landsat data of the epochs 1975, 1990, 2000, and 2014 | EU Science Hub. Publications Office of the European Union; 2016. Available: https://ec.europa.eu/jrc/en/publication/operating-procedure-production-global-human-settlement-layer-landsat-data-epochs-1975-1990-2000-and

50. U.S. Department of State - Humanitarian Information Unit. Global LSIB Polygons Detailed - Humanitarian Data Exchange. Humanitarian Data Exchange (HDX); Available: https://data.humdata.org/dataset/global-lsib-polygons-detailed

51. Exum NG, Olórtegui MP, Yori PP, Davis MF, Heaney CD, Kosek M, et al. Floors and Toilets: Association of Floors and Sanitation Practices with Fecal Contamination in Peruvian Amazon Peri-Urban Households. Environmental Science & Technology. 2016;50: 7373–7381. doi:10.1021/acs.est.6b01283

52. Cattaneo M, Galiani S, Gertler P, Martinez S, Titiunik R. Housing, Health and Happiness. CEDLAS, Working Papers. 2008 [cited 25 Apr 2019]. Available: https://ideas.repec.org/p/dls/wpaper/0074.html

53. Badr HS, Colston JM, Nguyen N-LH, Chen YT, Ali SA, Rayamajhi A, et al. Spatiotemporal variation in risk of Shigella infection in childhood: a global risk mapping and prediction model using individual participant data. medRxiv; 2022. p. 2022.08.04.22277641. doi:10.1101/2022.08.04.22277641

54. Getachew A, Tadie A, G.Hiwot M, Guadu T, Haile D, G.Cherkos T, et al. Environmental factors of diarrhea prevalence among under five children in rural area of North Gondar zone, Ethiopia. Italian Journal of Pediatrics. 2018;44: 95. doi:10.1186/s13052-018-0540-7

55. Paul P. Socio-demographic and environmental factors associated with diarrhoeal disease among children under five in India. BMC Public Health. 2020;20: 1886. doi:10.1186/s12889-020-09981-y

56. Tusting LS, Gething PW, Gibson HS, Greenwood B, Knudsen J, Lindsay SW, et al. Housing and child health in sub-Saharan Africa: A cross-sectional analysis. PLoS Med. 2020;17: e1003055. doi:10.1371/journal.pmed.1003055

57. Tusting LS, Bottomley C, Gibson H, Kleinschmidt I, Tatem AJ, Lindsay SW, et al. Housing Improvements and Malaria Risk in Sub-Saharan Africa: A Multi-Country Analysis of Survey Data. von Seidlein L, editor. PLOS Medicine. 2017;14: e1002234. doi:10.1371/journal.pmed.1002234

58. Aftab A, Noor A, Aslam M. Housing quality and its impact on Acute Respiratory Infection (ARI) symptoms among children in Punjab, Pakistan. PLOS Glob Public Health. 2022;2: e0000949. doi:10.1371/journal.pgph.0000949

59. Mengersen K, Morawska L, Wang H, Murphy N, Tayphasavanh F, Darasavong K, et al. The effect of housing characteristics and occupant activities on the respiratory health of women and children in Lao PDR. Sci Total Environ. 2011;409: 1378–1384. doi:10.1016/j.scitotenv.2011.01.016

60. Islam F, Sarma R, Debroy A, Kar S, Pal R. Profiling Acute Respiratory Tract Infections in Children from Assam, India. J Glob Infect Dis. 2013;5: 8–14. doi:10.4103/0974-777X.107167

61. Akinyemi JO, Morakinyo OM. Household environment and symptoms of childhood acute respiratory tract infections in Nigeria, 2003-2013: a decade of progress and stagnation. BMC Infect Dis. 2018;18: 296. doi:10.1186/s12879-018-3207-5

62. Furnival-Adams J, Olanga EA, Napier M, Garner P. House modifications for preventing malaria. Cochrane Database Syst Rev. 2020;10: CD013398. doi:10.1002/14651858.CD013398.pub2

63. Mshamu S, Mmbando A, Meta J, Bradley J, Bøjstrup TC, Day NPJ, et al. Assessing the impact of a novel house design on the incidence of malaria in children in rural Africa: study protocol for a household-cluster randomized controlled superiority trial. Trials. 2022;23: 519. doi:10.1186/s13063-022-06461-z

64. Lwetoijera DW, Kiware SS, Mageni ZD, Dongus S, Harris C, Devine GJ, et al. A need for better housing to further reduce indoor malaria transmission in areas with high bed net coverage. Parasites Vectors. 2013;6: 57. doi:10.1186/1756-3305-6-57

65. Kua KP, Lee SWH. Randomized trials of housing interventions to prevent malaria and Aedes-transmitted diseases: A systematic review and meta-analysis. PLoS One. 2021;16: e0244284. doi:10.1371/journal.pone.0244284

66. Msugupakulya BJ, Kaindoa EW, Ngowo HS, Kihonda JM, Kahamba NF, Msaky DS, et al. Preferred resting surfaces of dominant malaria vectors inside different house types in rural south-eastern Tanzania. Malar J. 2020;19: 22. doi:10.1186/s12936-020-3108-0

67. Lindsay SW, Jawara M, Paine K, Pinder M, Walraven GEL, Emerson PM. Changes in house design reduce exposure to malaria mosquitoes. Tropical Medicine & International Health. 2003;8: 512–517. doi:10.1046/j.1365-3156.2003.01059.x

68. Jatta E, Jawara M, Bradley J, Jeffries D, Kandeh B, Knudsen JB, et al. How house design affects malaria mosquito density, temperature, and relative humidity: an experimental study in rural Gambia. The Lancet Planetary Health. 2018;2: e498–e508. doi:10.1016/S2542-5196(18)30234-1

69. Kirby MJ, West P, Green C, Jasseh M, Lindsay SW. Risk factors for house-entry by culicine mosquitoes in a rural town and satellite villages in The Gambia. Parasites & Vectors. 2008;1: 41. doi:10.1186/1756-3305-1-41

70. Lindsay SW, Jawara M, Mwesigwa J, Achan J, Bayoh N, Bradley J, et al. Reduced mosquito survival in metal-roof houses may contribute to a decline in malaria transmission in sub-Saharan Africa. Sci Rep. 2019;9: 7770. doi:10.1038/s41598-019-43816-0

71. Snyman K, Mwangwa F, Bigira V, Kapisi J, Clark TD, Osterbauer B, et al. Poor Housing Construction Associated with Increased Malaria Incidence in a Cohort of Young Ugandan Children. The American Journal of Tropical Medicine and Hygiene. 2015;92: 1207–1213. doi:10.4269/ajtmh.14-0828

72. Rek JC, Alegana V, Arinaitwe E, Cameron E, Kamya MR, Katureebe A, et al. Rapid improvements to rural Ugandan housing and their association with malaria from intense to reduced transmission: a cohort study. The Lancet Planetary Health. 2018;2: e83–e94. doi:10.1016/S2542-5196(18)30010-X

73. Okiring J, Olwoch P, Kakuru A, Okou J, Ochokoru H, Ochieng TA, et al. Household and maternal risk factors for malaria in pregnancy in a highly endemic area of Uganda: a prospective cohort study. Malaria Journal. 2019;18: 144. doi:10.1186/s12936-019-2779-x

74. Yé Y, Hoshen M, Louis V, Séraphin S, Traoré I, Sauerborn R. Housing conditions and Plasmodium falciparum infection: protective effect of iron-sheet roofed houses. Malar J. 2006;5: 8. doi:10.1186/1475-2875-5-8

75. Ghebreyesus TA, Haile M, Witten KH, Getachew A, Yohannes M, Lindsay SW, et al. Household risk factors for malaria among children in the Ethiopian highlands. Transactions of the Royal Society of Tropical Medicine and Hygiene. 2000;94: 17–21. doi:10.1016/S0035-9203(00)90424-3

76. Hiscox A, Khammanithong P, Kaul S, Sananikhom P, Luthi R, Hill N, et al. Risk factors for mosquito house entry in the Lao PDR. PLoS One. 2013;8: e62769. doi:10.1371/journal.pone.0062769

77. Wolff CG, Schroeder DG, Young MW. Effect of improved housing on illness in children under 5 years old in northern Malawi: cross sectional study. BMJ. 2001;322: 1209– 1212. doi:10.1136/bmj.322.7296.1209

78. Coleman M, Coleman M, Mabaso MLH, Mabuza AM, Kok G, Coetzee M, et al. Household and microeconomic factors associated with malaria in Mpumalanga, South Africa. Transactions of The Royal Society of Tropical Medicine and Hygiene. 2010;104: 143–147. doi:10.1016/j.trstmh.2009.07.010

79. Liu JX, Bousema T, Zelman B, Gesase S, Hashim R, Maxwell C, et al. Is Housing Quality Associated with Malaria Incidence among Young Children and Mosquito Vector Numbers? Evidence from Korogwe, Tanzania. PLOS ONE. 2014;9: e87358. doi:10.1371/journal.pone.0087358

80. Tusting LS, Ippolito MM, Willey BA, Kleinschmidt I, Dorsey G, Gosling RD, et al. The evidence for improving housing to reduce malaria: a systematic review and meta-analysis. Malar J. 2015;14: 209. doi:10.1186/s12936-015-0724-1

81. Calderon-Anyosa R, Galvez-Petzoldt C, Garcia PJ, Carcamo CP. Housing Characteristics and Leishmaniasis: A Systematic Review. Am J Trop Med Hyg. 2018;99: 1547–1554. doi:10.4269/ajtmh.18-0037

82. Peña-García VH, Gómez-Palacio AM, Triana-Chávez O, Mejía-Jaramillo AM. Eco-Epidemiology of Chagas Disease in an Endemic Area of Colombia: Risk Factor Estimation, Trypanosoma cruzi Characterization and Identification of Blood-Meal Sources in Bugs. Am J Trop Med Hyg. 2014;91: 1116–1124. doi:10.4269/ajtmh.14-0112

83. Bustamante DM, Monroy C, Pineda S, Rodas A, Castro X, Ayala V, et al. Risk factors for intradomiciliary infestation by the Chagas disease vector Triatoma dimidiatain Jutiapa, Guatemala. Cad Saúde Pública. 2009;25: S83–S92. doi:10.1590/S0102-311X2009001300008

84. Rojas de Arias A, Ferro EA, Ferreira ME, Simancas LC. Chagas disease vector control through different intervention modalities in endemic localities of Paraguay. Bull World Health Organ. 1999;77: 331–339.

